# Dissecting the shared genetic architecture of suicide attempt, psychiatric disorders and known risk factors

**DOI:** 10.1101/2020.12.01.20241281

**Authors:** Niamh Mullins, Jooeun Kang, Adrian I Campos, Jonathan R I Coleman, Alexis C Edwards, Hanga Galfalvy, Daniel F Levey, Adriana Lori, Andrey Shabalin, Anna Starnawska, Mei-Hsin Su, Hunna J Watson, Mark Adams, Swapnil Awasthi, Michael Gandal, Jonathan D Hafferty, Akitoyo Hishimoto, Minsoo Kim, Satoshi Okazaki, Ikuo Otsuka, Stephan Ripke, Erin B Ware, Andrew W Bergen, Wade H Berrettini, Martin Bohus, Harry Brandt, Xiao Chang, Wei J Chen, Hsi-Chung Chen, Steven Crawford, Scott Crow, Emily DiBlasi, Philibert Duriez, Fernando Fernández-Aranda, Manfred M Fichter, Steven Gallinger, Stephen J Glatt, Philip Gorwood, Yiran Guo, Hakon Hakonarson, Katherine A Halmi, Hai-Gwo Hwu, Sonia Jain, Stéphane Jamain, Susana Jiménez-Murcia, Craig Johnson, Allan S Kaplan, Walter H Kaye, Pamela K Keel, James L Kennedy, Kelly L Klump, Robert D Levitan, Dong Li, Shih-Cheng Liao, Klaus Lieb, Lisa Lilenfeld, Chih-Min Liu, Pierre J Magistretti, Christian R Marshall, James E Mitchell, Eric T Monson, Richard M Myers, Dalila Pinto, Abigail Powers, Nicolas Ramoz, Stefan Roepke, Alessandro Rotondo, Vsevolod Rozanov, Stephen W Scherer, Christian Schmahl, Marcus Sokolowski, Michael Strober, Laura M Thornton, Janet Treasure, Ming T Tsuang, Maria C La Via, Stephanie H Witt, D Blake Woodside, Zeynep Yilmaz, Lea Zillich, Rolf Adolfsson, Ingrid Agartz, Tracy M Air, Martin Alda, Lars Alfredsson, Ole A Andreassen, Adebayo Anjorin, Vivek Appadurai, María Soler Artigas, Sandra Van der Auwera, M Helena Azevedo, Nicholas Bass, Claiton HD Bau, Bernhard T Baune, Frank Bellivier, Klaus Berger, Joanna M Biernacka, Tim B Bigdeli, Elisabeth B Binder, Michael Boehnke, Marco Boks, Rosa Bosch, David L Braff, Richard Bryant, Monika Budde, Enda M Byrne, Wiepke Cahn, Miguel Casas, Enrique Castelao, Jorge A Cervilla, Boris Chaumette, Sven Cichon, Aiden Corvin, Nicholas Craddock, David Craig, Franziska Degenhardt, Srdjan Djurovic, Howard J Edenberg, Ayman H Fanous, Jerome C Foo, Andreas J Forstner, Mark Frye, Janice M Fullerton, Justine M Gatt, Pablo V Gejman, Ina Giegling, Hans J Grabe, Melissa J Green, Eugenio H Grevet, Maria Grigoroiu-Serbanescu, Blanca Gutierrez, Jose Guzman-Parra, Steven P Hamilton, Marian L Hamshere, Annette Hartmann, Joanna Hauser, Stefanie Heilmann-Heimbach, Per Hoffmann, Marcus Ising, Ian Jones, Lisa A Jones, Lina Jonsson, René S Kahn, John R Kelsoe, Kenneth S Kendler, Stefan Kloiber, Karestan C Koenen, Manolis Kogevinas, Bettina Konte, Marie-Odile Krebs, Mikael Landén, Jacob Lawrence, Marion Leboyer, Phil H Lee, Douglas F Levinson, Calwing Liao, Jolanta Lissowska, Susanne Lucae, Fermin Mayoral, Susan L McElroy, Patrick McGrath, Peter McGuffin, Andrew McQuillin, Sarah Medland, Divya Mehta, Ingrid Melle, Yuri Milaneschi, Philip B Mitchell, Esther Molina, Gunnar Morken, Preben Bo Mortensen, Bertram Müller-Myhsok, Caroline Nievergelt, Vishwajit Nimgaonkar, Markus M Nöthen, Michael C O’Donovan, Roel A Ophoff, Michael J Owen, Carlos Pato, Michele T Pato, Brenda WJH Penninx, Jonathan Pimm, Giorgio Pistis, James B Potash, Robert A Power, Martin Preisig, Digby Quested, Josep Antoni Ramos-Quiroga, Andreas Reif, Marta Ribasés, Vanesa Richarte, Marcella Rietschel, Margarita Rivera, Andrea Roberts, Gloria Roberts, Guy A Rouleau, Diego L Rovaris, Dan Rujescu, Cristina Sánchez-Mora, Alan R Sanders, Peter R Schofield, Thomas G Schulze, Laura J Scott, Alessandro Serretti, Jianxin Shi, Stanley I Shyn, Lea Sirignano, Pamela Sklar, Olav B Smeland, Jordan W Smoller, Edmund J S Sonuga-Barke, Gianfranco Spalletta, John S Strauss, Beata Świątkowska, Maciej Trzaskowski, Gustavo Turecki, Laura Vilar-Ribó, John B Vincent, Henry Völzke, James TR Walters, Cynthia Shannon Weickert, Thomas W Weickert, Myrna M Weissman, Leanne M Williams, Major Depressive Disorder Working Group of the PsychiatricGenomics Consortium, Bipolar Disorder Working Group of the Psychiatric Genomics Consortium, Eating Disorders Working Group of the Psychiatric Genomics Consortium, German Borderline Genomics Consortium, Naomi R Wray, Clement Zai, Esben Agerbo, Anders D Børglum, Gerome Breen, Annette Erlangsen, Tõnu Esko, Joel Gelernter, David M Hougaard, Ronald C Kessler, Henry R Kranzler, Qingqin S Li, Nicholas G Martin, Andrew M McIntosh, Sarah E Medland, Ole Mors, Merete Nordentoft, Catherine M Olsen, David Porteous, Robert J Ursano, Danuta Wasserman, Thomas Werge, David C Whiteman, Cynthia M Bulik, Hilary Coon, Ditte Demontis, Anna R Docherty, Po-Hsiu Kuo, Cathryn M Lewis, J John Mann, Miguel E Rentería, Daniel J Smith, Eli A Stahl, Murray B Stein, Fabian Streit, Virginia Willour, Douglas M Ruderfer

## Abstract

Suicide is a leading cause of death worldwide and non-fatal suicide attempts, which occur far more frequently, are a major source of disability and social and economic burden. Both are known to have a substantial genetic etiology, which is partially shared and partially distinct from that of related psychiatric disorders. We conducted a genome-wide association study (GWAS) of 29,782 suicide attempt (SA) cases and 519,961 controls in the International Suicide Genetics Consortium and conditioned the results on psychiatric disorders using GWAS summary statistics, to investigate their shared and divergent genetic architectures. Two loci reached genome-wide significance for SA: the major histocompatibility complex and an intergenic locus on chromosome 7, which remained associated after conditioning and has previously been implicated in risk-taking, smoking, and insomnia. SA showed strong genetic correlation with psychiatric disorders, particularly major depression, and also with smoking, lower socioeconomic status, pain, lower educational attainment, reproductive traits, risk-taking, sleep disturbances, and poorer overall general health. After conditioning, the genetic correlations between SA and psychiatric disorders decreased, whereas those with non-psychiatric traits remained largely unchanged. Our results identify a risk locus that contributes more strongly to SA than other phenotypes and suggest the existence of a shared genetic etiology between SA and known risk factors that is not mediated by psychiatric disorders.

## Introduction

Suicide is a worldwide public health problem, accounting for close to 800,000 deaths per year^1^. Suicide attempt (SA), defined as non-fatal self-injurious behavior with the intent to die, has been estimated to occur over 20 times more frequently and is a major source of disability, reduced quality of life, and social and economic burden^1,2^. The lifetime prevalence of SA in adults ranges from 0.5-5% worldwide^3^. There are several well established comorbidities and risk factors for SA, with the presence of a psychiatric disorder having the strongest effect on lifetime suicide rates^4,5^. However, the vast majority of patients with psychiatric disorders never attempt suicide^6–8^. Other major risk factors for SA include prior self-injurious thoughts and behaviors^9^, physical illness or disability^10,11^, sleep disorders^12–15^, family history of psychiatric disorders^16^, substance abuse^17^, smoking^18–20^, impulsivity^21^ and social factors such as childhood maltreatment^21^, isolation^22^, and stressful life events^23^.

Both suicide and SA are moderately heritable, with estimates from genetic epidemiology studies in the range of 17-55%^24–26^. Several genome-wide association studies (GWAS) of SA have reported significant SNP-heritability estimates of ∼4%, pointing to an underlying polygenic architecture^27–31^. Using polygenic risk scoring or genetic correlation analyses, these studies have also demonstrated shared genetic etiology between SA and psychiatric disorders, with major depressive disorder (MDD) showing the largest genetic overlap^28,29,31^. This genetic overlap, along with the prevalence of MDD in the population^32^ make it a particularly salient risk factor. Importantly, genetic epidemiology studies have consistently indicated a genetic component of SA which is partially distinct from that of psychiatric disorders^25^. One GWAS of SA adjusted for the presence of a psychiatric disorder and estimated a SNP heritability of 1.9%^27^, suggesting that the genetic etiology of SA is likely to comprise genetic variants which confer risk more strongly to SA than psychiatric disorders, as well as variants that confer risk more strongly to psychiatric disorders than SA.

Few genetic samples have been collected specifically for SA, with studies often relying on individuals ascertained for psychiatric disorders. For example, a large GWAS of SA included over 6,500 cases from clinical cohorts of depression, bipolar disorder and schizophrenia cases, within the Psychiatric Genomics Consortium^31^. In a “SA within psychiatric diagnosis” study design, SA cases were compared with cases of the same psychiatric disorder without SA, in order to disentangle the genetic etiology of SA and psychiatric disorders. While GWAS of SA have found genome-wide significant associations^27–31^, thus far none have been replicated, possibly due to limited statistical power or different study designs which may probe varying components of the genetic etiology of SA. Depending on the method of ascertainment, the prevalence of psychiatric disorders may be much higher in SA cases than controls in these studies, which may confound the genetics of SA. Well-powered and carefully designed studies are necessary to advance our understanding of the genetics of SA and dissect the contribution of genetic variation to SA versus psychiatric disorders.

Here, we present the first collaborative GWAS meta-analysis of SA from the International Suicide Genetics Consortium, including over 29,000 cases of suicide or SA from 15 institutes or consortia worldwide. We identify novel loci implicated in SA, disentangle the genetic etiology of SA from that of MDD and psychiatric disorders and characterize the genetic relationship between SA, psychiatric disorders, and a range of known risk factors.

## Methods

### Cohorts and case definition

This study included 21 cohorts worldwide, many of which have been described previously (Table S1, Supplementary Note). These included cohorts ascertained for psychiatric disorders, including substance use (15 cohorts), studies of suicide or SA (4 cohorts), and population-based biobanks (2 cohorts). Cases were individuals who died by suicide (2 cohorts) or made a non-fatal suicide attempt (19 cohorts). A non-fatal suicide attempt was defined as a lifetime act of deliberate self-harm with intent to result in death. Information on SA was ascertained using structured clinical interviews for 15 cohorts, self-report questionnaires for 2 cohorts, and hospital records or International Classification of Diseases (ICD) codes for 2 cohorts. Cases of death by suicide (2 cohorts) were ascertained from the Utah State Office of the Medical Examiner or the Medical Examiner’s Office of the Hyogo Prefecture and the Division of Legal Medicine, at the Kobe University Graduate School of Medicine in Japan. A proportion of cases from the iPSYCH and Columbia University cohorts were also individuals who had died by suicide, determined using the Cause of Death Register in Denmark and The Columbia Classification Algorithm for Suicide Assessment respectively^33^. Individuals endorsing suicidal ideation only were not included as cases. There were 14 cohorts of European ancestry, 2 cohorts of admixed African American ancestry, and 5 cohorts of East Asian ancestry. All individual studies received institutional and ethical approval from their local institutional review board (Table S1). Detailed information on the ascertainment and case definition for each cohort is included in the Supplementary Note. Supplementary Table 1 contains an overview of cohort characteristics.

### Control definition

For the primary GWAS, controls included all individuals with no evidence of SA, including those ascertained for having a psychiatric disorder. Controls from the general population were screened for the absence of SA if such information was available; however since the prevalence of SA in the general population is low (∼2%)^3^, some cohorts included unscreened controls. Amongst controls ascertained for having a psychiatric disorder, all were screened for the absence of lifetime SA. Controls from the general population were not screened for the absence of psychiatric disorders and no controls were screened for suicidal ideation. A GWAS of SA within psychiatric diagnosis was also conducted, where controls were individuals with the same psychiatric disorder as the SA cases in each cohort, and were all screened for the absence of lifetime SA. Cohorts were included in the GWAS of SA in the general population and/or the GWAS of SA within psychiatric diagnosis, depending on the characteristics of the controls available, and therefore there is some overlap of individuals and cohorts between the GWAS. The primary GWAS of SA included 29,782 cases and 519,961 controls from 18 cohorts and the GWAS of SA within psychiatric diagnosis included 14,847 cases and 69,951 controls from 13 cohorts (Table 1).

**Table 1.** Numbers of cases and controls for 21 cohorts in the International Suicide Genetics Consortium.

| **Cohort** | **SA\*** |  | **SA within psychiatric diagnosis\*** |  |
| --- | --- | --- | --- | --- |
|  | **Cases** | **Controls** | **Cases** | **Controls** |
| Psychiatric Genomics Consortium MDD | 1,528 | 16,626 | 1,677 | 8,793 |
| Psychiatric Genomics Consortium BIP | 3,214 | 17,642 | 3,214 | 5,408 |
| Psychiatric Genomics Consortium SCZ | 1,640 | 7,112 | 1,668 | 2,928 |
| Psychiatric Genomics Consortium ED | 170 | 5,070 | 198 | 583 |
| Army STARRs | 670 | 10,637 | 376 | 3,447 |
| German Borderline Genomics Consortium | 481 | 1,653 | 481 | 144 |
| UK Biobank | 2,433 | 334,766 | 2,149 | 35,912 |
| iPSYCH | 7,003 | 52,227 | - | - |
| Janssen | 255 | 1,684 | - | - |
| Yale-Penn | 475 | 1,817 | - | - |
| GISS Ukraine | 660 | 660 | - | - |
| Columbia University | 577 | 1,233 | - | - |
| Australian Genetics of Depression Study and QSkin Study | 2,792 | 20,193 | 2,792 | 8,718 |
| University of Utah | 4,692 | 20,702 | - | - |
| Japan (EAS) | 746 | 14,049 | - | - |
| CONVERGE (EAS) | 1,148 | 6,515 | 1,148 | 1,183 |
| Taiwan MDD (EAS) | - | - | 222 | 318 |
| Taiwan BIP (EAS) | - | - | 235 | 397 |
| Taiwan SCZ (EAS) | - | - | 332 | 1,004 |
| Grady Trauma Project (admixed AA) | 669 | 4,473 | 355 | 1,116 |
| Yale-Penn (admixed AA) | 629 | 2,902 | - | - |
| **Total** | **29,782** | **519,961** | **14,847** | **69,951** |

SA - suicide attempt, MDD - major depressive disorder, BIP - bipolar disorder, SCZ - schizophrenia, ED - eating disorder, EAS - East Asian ancestry, AA - African American ancestry. All other samples are of European ancestries. \*GWAS contain some overlapping individuals and cohorts.

### Genotyping, quality control and imputation

Cohorts were required to have a minimum of 200 cases prior to quality control for inclusion in the GWAS meta-analysis. Samples underwent standard genotyping, quality control and imputation, according to the local protocol for each study. Briefly, samples were genotyped on microarrays with the exception of one study (CONVERGE) that used low-coverage sequencing. Parameters used to retain individuals and SNPs after quality control for missingness, relatedness and Hardy-Weinberg equilibrium are outlined in the Supplementary Note. Imputation was performed using the appropriate ancestry reference panels, resulting in > 7.7 million SNPs that were well-represented across cohorts. Full details of the genotyping, quality control and imputation for each cohort are available in the Supplementary Note. Identical individuals between the Psychiatric Genomics Consortium (PGC) and UK Biobank cohorts were detected using genotype-based checksums (https://personal.broadinstitute.org/sripke/share_links/zpXkV8INxUg9bayDpLToG4g58TMtjN_PGC_SCZ_w3.0718d.76) and removed from PGC cohorts. There was no other known overlap of controls remaining between any of the 21 cohorts after QC.

### Genome-wide association study

GWAS were performed in each cohort separately by the collaborating research team and analysis procedures are outlined in the Supplementary Note. GWAS were conducted within ancestry group, covarying for genetic ancestry-informative principal components (PCs), genomic relatedness matrices or factors capturing site of recruitment or genotyping batch, as required. The LD Score regression (LDSC) intercept was calculated for all GWAS results to assess potential confounding from cryptic relatedness or population stratification^34^. For any studies with a significant intercept (P<0.05), the GWAS summary statistics were corrected for confounding by multiplying the standard error per SNP by the square root of the LDSC intercept^34^. A meta-analysis of GWAS summary statistics was conducted using an inverse variance-weighted fixed effects model (standard error) in METAL^35^, implemented using the Rapid Imputation for COnsortias PIpeLIne (RICOPILI)^36^, for the GWAS of SA in the general population, and the GWAS of SA within psychiatric diagnosis. The meta-analyses were performed across all cohorts regardless of ancestry. The weighted mean allele frequency and imputation INFO score per SNP was calculated, weighted by the effective sample size per cohort. SNPs with a weighted minor allele frequency of < 1%, weighted imputation INFO score < 0.6 or SNPs present in < 80% of total effective sample size were removed from the meta-analysis results. A genome-wide significant locus was defined as the region around a SNP with P<5.0×10^−8^ with linkage disequilibrium (LD) r^2^ > 0.1, within a 3,000 kilobase (kb) window, based on the LD structure of the Haplotype Reference Consortium (HRC) European ancestry reference panel v1.0^37^.

### mtCOJO

The results of the GWAS of SA were conditioned on the genetics of MDD using mtCOJO (multi-trait-based conditional & joint analysis using GWAS summary data)^38^, implemented in GCTA software^39^. mtCOJO^38^ estimates the effect size of a SNP on an outcome trait (eg. SA) conditioned on exposure trait(s) (eg. MDD). It first uses the genome-wide significant SNPs for the exposure trait as instruments to estimate the effect of the exposure on the outcome, and then performs a genome-wide conditioning of the estimated effect from the exposure, resulting in conditioned effect sizes and P values for the outcome trait. We conditioned SA on MDD, since MDD is the most prevalent psychiatric disorder among individuals who die by suicide^40^ and has the highest genetic correlation with SA among psychiatric disorders (rg=0.44)^28^. mtCOJO analysis was performed on the SA as the outcome trait. For this, GWAS summary statistics from the European-only subset of the SA meta-analysis were used (26,590 cases and 492,022 controls), since mtCOJO requires an ancestry-matched LD reference panel. The PGC MDD GWAS summary statistics (excluding 23andMe)^41^ were used for the exposure trait. mtCOJO is robust to overlap in samples contributing to the GWAS of the exposure and outcome. In the selection of SNPs as instruments, independence was defined as SNPs more than 1 megabase (Mb) apart or with an LD r^2^ value < 0.05 based on the 1000 Genomes Project Phase 3 European reference panel^42^. To obtain at least 10 independent instruments for MDD, the genome-wide significance threshold was adjusted to P<5.0×10^−7^, leading to 15 SNPs used. In a further sensitivity analysis, GWAS summary statistics for bipolar disorder (BIP)^43^ and schizophrenia (SCZ)^44^ were additionally included as exposure traits.

### LD Score regression (LDSC)

LDSC^34^ was used to estimate the phenotypic variance in SA explained by common SNPs (SNP-heritability, 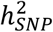) from GWAS summary statistics. 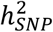 was calculated on the liability scale assuming a lifetime prevalence of SA in the general population of 2%, which is the middle of the range reported worldwide^3^. For the GWAS of SA within psychiatric diagnosis, 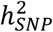 was calculated on the liability scale using a prevalence of SA in psychiatric populations ranging from 10-20%. LDSC bivariate genetic correlations attributable to genome-wide SNPs (rg) were estimated between all GWAS of SA and between each GWAS of SA and a range of psychiatric disorders, self-harm ideation and propensity towards risk-taking behavior (risk tolerance), using the largest available GWAS summary statistics (Table S11). The Bonferroni corrected significance threshold was P<0.0042, adjusting for 12 traits tested. The difference between the rg of SA before and after conditioning on MDD was tested for deviation from 0, using the block jackknife method, implemented by the LDSC software^45^. The rg of each SA GWAS with 768 other non-overlapping human diseases and traits was calculated on LD Hub (http://ldsc.broadinstitute.org)^46^ (Bonferroni corrected significance threshold P<6.51×10^−5^ for each GWAS). Before analysis, traits were categorized manually into risk factor groups previously ascribed to SA^4,5,10^: autoimmune disease, neurologic disease, heart disease, hypertension, diabetes, kidney disease, cancer, alcohol use, smoking, pain, psychiatric, sleep, life stressors, socioeconomic, and education/cognition (Table S12). A second reviewer validated the categories assigned to traits and their relevance to SA risk. Overlapping traits were appended.

### Gene-based, gene-set and tissue-set enrichment analyses

P values quantifying the degree of association of genes and gene-sets with SA based on the GWAS of SA in the general population were generated using MAGMA (v1.08), implemented in FUMA (v1.3.6a) (https://fuma.ctglab.nl)^47,48^. Gene-based tests were performed for 18,517 genes and a Bonferroni correction was applied for the number of genes tested (P<2.70×10^−6^). A total of 11,638 curated gene sets from MSigDB V7.0 were also tested for association with SA (Bonferroni-corrected significance threshold P<4.30×10^−6^). Competitive gene-set tests were conducted correcting for gene size, variant density and LD within and between genes. Gene-sets including < 10 genes were excluded. Finally, tissue-set enrichment analyses were performed using MAGMA^48^ implemented in FUMA^47^, to test for enrichment of genetic associations with SA in genes expressed in 54 tissues from the Genotype-Tissue Expression (GTEx) project V8^49^. The significance threshold was P<9.26×10^−4^, adjusting for the number of tissues tested.

### Integrative eQTL analysis

A transcriptome-wide association study (TWAS) was conducted using FUSION software^50^ and precomputed expression reference weights from PsychENCODE data^51^. The PsychENCODE Consortium has conducted a genome-wide eQTL analysis using 1,321 brain samples, predominantly from the dorsolateral prefrontal cortex^51^. For genes with significant *cis*-SNP heritability (13,435 genes), a TWAS was performed to test whether SNPs influencing brain gene expression are also associated with SA, using the meta-analysis results from the GWAS of SA in the general population (Bonferroni corrected significance threshold P<4.28×10^−6^).

### Polygenic risk scoring analysis

Polygenic risk scores (PRS) for SA were tested for association with SA or death by suicide in independent target cohorts. The target cohorts used were PGC MDD, PGC BIP, PGC SCZ, CONVERGE (a cohort of East Asian ancestry), and the University of Utah cohort (a sample of individuals who died by suicide). The meta-analysis of SA was repeated excluding each of these cohorts in turn, to create independent discovery and target datasets. PRS were tested for association with SA versus controls in all five of the target samples. PRS were also tested for association with SA within psychiatric diagnosis in the PGC MDD, BIP and SCZ samples. Analyses in the PGC datasets were repeated using the PRS for SA in the general population generated from the GWAS results after conditioning on MDD. The Bonferroni corrected significance threshold is P<3.57×10^−3^, correcting for 14 tests. The analyses performed are summarized in Table S2.

PRS analyses were performed using PRS-CS^52^, a method which uses a Bayesian regression framework and places a continuous shrinkage prior on the effect sizes of all SNPs in the discovery GWAS summary statistics. Continuous shrinkage priors allow a specific amount of shrinkage to be applied to each SNP, which is adaptive to the strength of its association signal in the discovery GWAS and the LD structure from an external reference panel^52^. The 1000 Genomes European or East Asian reference panels^42^, as appropriate, were used to estimate LD between SNPs. PRS were calculated for each individual in the target cohorts using standard procedures. PLINK 1.9^53^ was used to weight all SNPs by their effect sizes calculated using PRS-CS and sum all SNPs into PRS for each individual in the target cohort. PRS were tested for association with case versus control status in the target cohort using a logistic regression model, covarying for PCs, genomic relatedness matrices or factors capturing site of recruitment or batch effects, as required. The amount of phenotypic variance explained by the PRS (R^2^) was calculated on the liability scale, which accounts for the proportion of cases in the target sample and the proportion of cases in the population^54^. Calculations assumed a lifetime prevalence of SA in the general population of 2%^3^ and a lifetime prevalence of SA in MDD, BIP, and SCZ of 16%, 37% and 36% respectively. These numbers represent the observed prevalence of SA in these disorders in the PGC cohorts.

### Pairwise GWAS

Pairwise GWAS^55^ was used to investigate genome-wide significant loci for SA and overlapping causal variants with propensity towards risk-taking behavior^56^ and lifetime smoking index^57^. These phenotypes were chosen because they share genome-wide significant loci in the same region as the genome-wide significant locus on chromosome 7 in the GWAS of SA and SA conditioned on MDD. The genome-wide significant locus on chromosome 6 is in the major histocompatibility complex and due to the complex long-range LD of this region, it was not included for this analysis. Pairwise GWAS uses association statistics from two GWAS to estimate the probability that a genomic region 1) contains a genetic variant that influences only the first trait, 2) contains a genetic variant that influences only the second trait, 3) contains a shared causal or pleiotropic variant, and 4) contains two independent variants in the same region, one influencing the first trait and the other influencing the second trait. The GWAS summary statistics from the European-only subset of the SA meta-analysis (26,590 cases and 492,022 controls) were used for Pairwise GWAS as the method requires an ancestry-matched LD reference panel. The genome was divided into approximately independent LD blocks based on patterns of LD in the European population in Phase 1 of the 1000 Genomes Project, as previously described^55^. We divided the 3 Mb-wide genome block containing the genome-wide significant locus for SA on chromosome 7 into two blocks to separate the two independent causal variants for risk-taking behavior in that region (rs8180817 and rs4275159, LD r^2^=0.001)^56^. The fgwas package^58^ was used to determine the baseline correlation between the two GWAS by extracting all genomic regions with a posterior probability of containing an association less than 0.2 and calculating the correlation in the Z-scores between the two GWAS. This summary statistic-level correlation was used as a correction factor to each Pairwise GWAS analysis.

## Results

### Study description and samples analyzed

We conducted a primary GWAS meta-analysis of SA (29,782 cases, 519,961 controls) from 18 cohorts (Table 1), which included both population-based and clinically ascertained samples for psychiatric disorders (see Methods). The majority (n=26,590) of cases were individuals of European ancestries but cases also included 1,894 individuals of East Asian ancestries and 1,298 individuals of admixed African American ancestries. Case definition was lifetime SA, with ∼20% (n=5,438) of cases having died by suicide (see Methods). To investigate the shared and divergent genetic architectures of SA and psychiatric disorders, we performed two additional analyses. We conditioned our primary GWAS results using GWAS summary statistics for MDD, to remove the genetic effects mediated by MDD, the most commonly comorbid psychiatric disorder with SA. Furthermore, we conducted a GWAS of SA versus no SA among individuals with a psychiatric diagnosis in 14,847 cases and 69,951 controls from 13 cohorts.

### SA shows significant SNP-heritability and polygenic risk association with death by suicide

In the primary GWAS of SA, 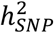 estimated using LDSC was 6.8% (SE=0.005, P=2.00×10^−42^) on the liability scale. The genomic inflation factor (λ_GC_) was 1.23, the LDSC intercept was 1.04 (SE=0.01, P=2.84×10^−4^) and the LDSC attenuation ratio was 0.14 (SE=0.04), indicating that the majority of inflation of the GWAS test statistics was due to polygenicity. PRS for SA were tested in five target SA cohorts, which were each excluded in turn from the discovery GWAS to ensure independent discovery and target samples (Table S2). SA PRS were significantly associated with SA in the PGC MDD, PGC BIP and PGC SCZ cohorts, with a phenotypic variance explained (R^2^) of 0.69% (P=7.17×10^−15^), 0.68% (P=8.11×10^−28^) and 0.88% (P=1.24×10^−^ 17) respectively, on the liability scale. PRS for SA were also associated with death by suicide in the University of Utah cohort, explaining slightly more phenotypic variance (R^2^=1.08%, P=9.79×10^−81^). The genetic correlation between the University of Utah GWAS of suicide death and SA from a meta-analysis of the remaining cohorts in our study was 0.77 (SE=0.08, P=1.54×10^−20^). Examining the performance of SA PRS across ancestry showed a significant association with SA in the CONVERGE East Asian cohort, although with a lower variance explained (R^2^=0.25%, P=3.06×10^−3^) (Table S2).

### GWAS of SA identifies locus with stronger effect on SA than psychiatric disorders

The GWAS of SA identified two genome-wide significant loci (P<5×10^−8^) (Figure 1a, Table S3). The locus most highly associated with SA was in an intergenic region on chromosome 7 (index SNP rs62474683, OR for A allele = 1.06 [1.04-1.08], P=1.91×10^−10^, frequency in SA cases = 0.52). The second genome-wide significant locus was in the major histocompatibility complex (MHC) (index SNP rs71557378, OR for T allele= 1.10 [1.06-1.13], P=1.97×10^−8^, frequency in SA cases = 0.91). After conditioning the genetic effects of SA (European-only subset) on the genetic effects of MDD using mtCOJO, only the chromosome 7 locus remained genome-wide significant (index SNP = rs62474683, OR for A allele = 1.06 [1.04-1.08], P=1.33×10^−^8, Figure 1a). In the GWAS of SA within psychiatric diagnosis, this index SNP had a slightly smaller effect size on SA (index SNP = rs62474683, OR for A allele = 1.04 [1.01-1.07], P=0.007), but no SNPs reached genome-wide significance in this analysis. Examining the intergenic locus on chromosome 7 in published GWAS results using Open Targets Genetics web portal^59^ (https://genetics.opentargets.org), showed smaller and non-significant effects on all psychiatric disorders tested (Figure 1b). However, the index SNP from our SA GWAS has been implicated at genome-wide significance in lifetime smoking index^57^ (which accounts for duration and amount of smoking), and propensity towards risk-taking behavior^56^, although with smaller effect sizes than on SA (Figure 1b, Table S4-5). Pairwise GWAS analysis on the genomic region containing the chromosome 7 locus indicated that the causal variant is most likely shared between SA and these phenotypes (lifetime smoking index: posterior probability = 0.997, risk-taking behavior: posterior probability = 1) (Table S13). Furthermore, a variant in high LD with the index SNP on chromosome 7 (rs12666306, LD r^2^=0.94) has a positive genome-wide significant effect on insomnia (reported in GWAS catalog, full summary statistics not available)(Figure 1b, Table S4-5). The index SNP for SA has also been implicated in self-harm ideation^60^, although with a smaller effect size than on SA (Figure 1b).

**Figure 1:**
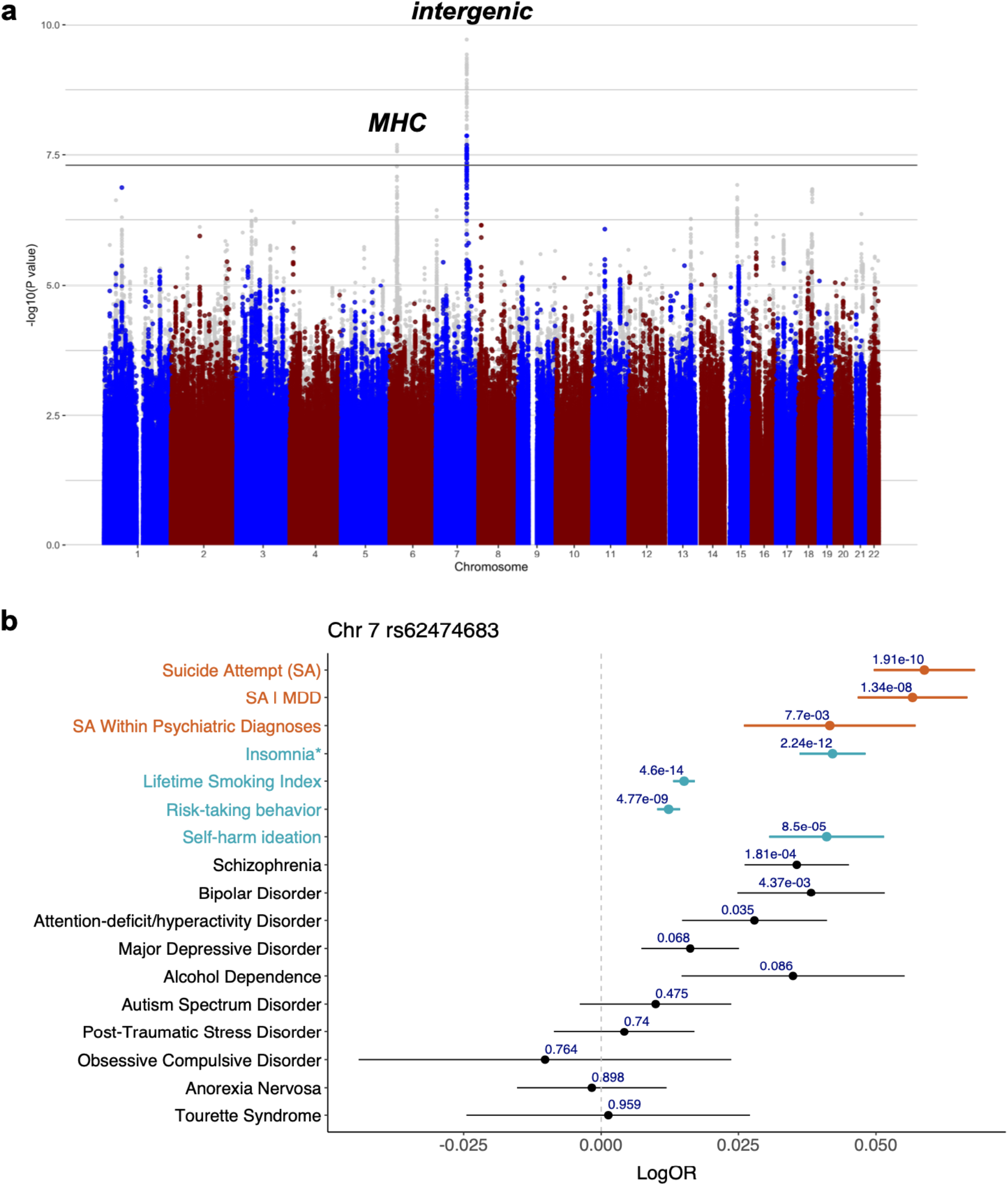
Genome-wide significant locus contributes to suicide attempt more strongly than psychiatric disorders and other traits. a) Manhattan plot: The *x-*axis shows genomic position and the *y-*axis shows statistical significance as – log10(P value). The grey points in the background depict the GWAS results for SA and the colored points in the foreground depict the results after conditioning SA on major depressive disorder (MDD), which was performed on the European meta-analysis results. The horizontal line shows the genome-wide significance threshold (P<5.0×10^−8^). b) Forest plot: The points indicate the log odds ratio of the A allele at rs62474683 (index SNP for SA on chromosome 7) on each phenotype and the error bars show the standard error. The P value of association with each phenotype is shown above the error bars. For insomnia, the effect size of a variant in high LD with the index SNP is shown instead (rs12666306 A allele, LD r^2^=0.94 with SA index SNP).

Enrichment analyses using MAGMA^48^ and the GWAS results for SA indicated significant enrichment of SA associations in 7 genes (Table S6), including *BTN2A1* which is a brain-expressed gene^61^ located within the MHC, that encodes a plasma membrane protein. There was no enrichment of SA association signal in any of the biological gene sets (Table S7) or in the set of genes expressed in any of the 54 GTEx tissues tested (Table S8). Examining individual genes, our transcriptome-wide association study (TWAS) found 5 genes for which SA risk alleles were significantly associated with brain gene expression: *ERC2, RP11*−*266A24*.*1, TIAF1, BACE2, NUFIP2* (P<4.28×10^−6^) (Table S9). None of the TWAS significant genes were located in genome-wide significant loci.

### Evidence for substantial proportion of SNP-heritability of SA not mediated by psychiatric disorders

We employed two approaches to assess the genetic architecture of SA after accounting for psychiatric disorders: 1) we statistically conditioned out genetic effects mediated by MDD and 2) we directly analyzed SA versus no SA among psychiatric disorder cases (see Methods). The statistical conditioning was performed on the European-only subset of the meta-analysis, in which the 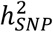 of SA was 7.5% (SE=0.006, P=3.02×10^−40^) on the liability scale (Table S10). Conditioning these SA GWAS results on MDD resulted in a 45% decrease in the 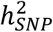 of SA to 4.1% (SE=0.005, P=1.20×10^−16^) on the liability scale (Table S10). This conditioned estimate was comparable with estimates of the 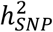 of SA within psychiatric diagnosis, which ranged from 3.7% to 4.6%, using a prevalence of SA in psychiatric populations from 10-20% (P<1.35×10^−3^). Conditioning SA on BIP and SCZ in addition to MDD did not change the 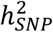 estimate 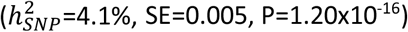.

The genetic correlation between the GWAS of SA and SA within psychiatric diagnosis was 0.93 (SE=0.09, P=5.35×10^−24^). PRS for SA were significantly associated with SA within psychiatric diagnosis in the PGC cohorts, with an R^2^ of 0.43% (P=5.83×10^−6^), 0.81% (P=2.33×10^−11^) and 0.71% (P=5.78×10^−6^) on the liability scale for SA in MDD, BIP and SCZ respectively (Table S2). After conditioning the GWAS of SA on MDD, the genetic correlation with the GWAS of SA within psychiatric diagnosis was not significantly different from 1 (rg=1.13, SE=0.13) (Table S10). After conditioning on MDD, PRS for SA remained significantly associated with SA within psychiatric diagnosis in the PGC cohorts, with slightly lower phenotypic variance explained (0.32%, 0.67% and 0.46% for SA in MDD, BIP and SCZ respectively), consistent with the reduction in 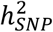 (Table S2).

### Significant genetic overlap between SA and psychiatric traits or disorders

Genetic correlations were calculated to explore the genetic overlap between SA and 12 psychiatric traits or disorders, before and after conditioning on MDD. SA showed a significant genetic correlation with 11 traits or disorders tested, most strongly with self-harm ideation (rg=0.81, SE=0.06, P=3.52×10^−36^) and MDD (rg=0.78, SE=0.03, P=5.82×10^−112^) (Figure 2, Table S11). Significant genetic correlations were also observed between SA and SCZ, attention-deficit/hyperactivity disorder (ADHD), BIP, post-traumatic stress disorder (PTSD) and alcohol dependency (rg=0.46-0.73) (Figure 2, Table S11).

**Figure 2:**
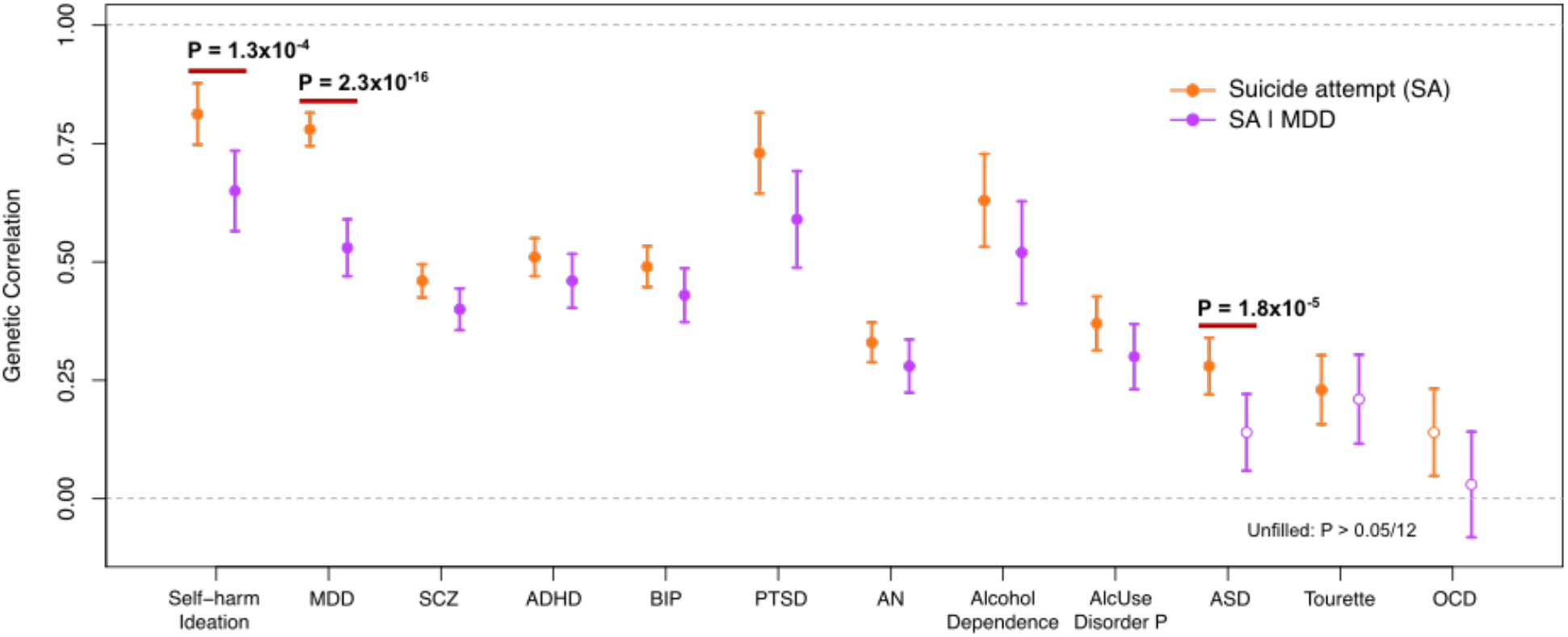
Substantial genetic correlation of suicide attempt with psychiatric traits or disorders before and after conditioning on major depressive disorder. Unfilled points indicate genetic correlations that did not pass the Bonferroni-corrected significance threshold P<4.17×10^−3^ (12 traits tested). Error bars represent the standard error. P values indicate significant differences in genetic correlation after conditioning, that pass the Bonferroni correction. SA|MDD-suicide attempt conditioned on major depressive disorder, MDD-major depressive disorder, SCZ-schizophrenia, ADHD-attention-deficit/hyperactivity disorder, BIP-bipolar disorder, PTSD-post-traumatic stress disorder, AN-anorexia nervosa, AlcUse Disorder P-Alcohol Use Disorders Identification Test-P (AUDIT-P, measure of problematic consequences of drinking), ASD-autism spectrum disorder, OCD-obsessive compulsive disorder.

To investigate whether these genetic correlations were mediated by the genetics of MDD, we estimated genetic correlations with the same traits and disorders after conditioning the GWAS of SA on MDD (SA|MDD). Genetic correlations with all psychiatric disorders remained significant after conditioning except for autism spectrum disorder (ASD) and Tourette syndrome (Figure 2, Table S11). As expected, the rg with MDD significantly decreased after conditioning (P=2.3×10^−16^ block jackknife), as well as the rg with self-harm ideation (P=1.3×10^−4^ block jackknife) and ASD (P=1.8×10^−5^ block jackknife) (Figure 2, Table S11). The remaining psychiatric disorders did not show significant differences in rg after conditioning on MDD, after Bonferroni correction. Since conditional analysis only removes SNP effects on SA mediated by MDD, the remaining genetic correlation between SA|MDD and MDD (rg=0.53, SE=0.06, P=8.9×10^−19^) indicates pleiotropic SNP effects.

Examining the genetic correlations between SA within psychiatric diagnosis and psychiatric disorders, most genetic correlations were comparable to those observed with SA|MDD (Table S11). Genetic correlations of SA within psychiatric diagnosis and MDD (rg=0.52, SE=0.11, P=4.48×10^−6^), ADHD (rg=0.60, SE=0.12, P=7.08×10^−7^), and PTSD (rg=0.56, SE=0.19, P=3.41×10^−3^) were significant after Bonferroni correction. As exceptions, BIP and SCZ had non-significant genetic correlations with SA within psychiatric diagnosis (SCZ: rg=-0.07, SE=0.075, P=3.24×10^−1^, BIP: rg=-0.08, SE=0.10, P=4.38×10^−1^). This is consistent with a previous report that BIP and SCZ cases who had attempted suicide did not have higher BIP or SCZ PRS, compared with cases who did not attempt suicide^31^.

### Substantial shared genetic architecture of SA and non-psychiatric risk factors not mediated by MDD

To assess the shared genetic architecture of SA, psychiatric, and non-psychiatric phenotypes, we calculated genetic correlations of our three GWAS (SA, SA|MDD and SA within psychiatric diagnosis) with 768 non-overlapping phenotypes^46^. We grouped 269 of these phenotypes into 15 categories of previously identified risk factors for SA^4,5,10^ (see Methods). There were 194 phenotypes which showed a significant rg with SA, 133 of which were in one of the pre-defined SA risk categories (Figure 3a, Table S12). The most significant genetic correlations were predominantly with traits related to depressive symptoms, smoking, and socioeconomic status. Examining phenotypes in the risk categories after conditioning on MDD, 81 phenotypes retained a significant genetic correlation with SA (Table S12). Within the psychiatric risk category, there was an average decrease in the magnitude of genetic correlation of 33% with SA after conditioning, whereas the genetic correlation values in other risk categories were much less affected by conditioning (smoking: 3% decrease, education/cognition: 0.74% increase, alcohol: 12.5% decrease, and socioeconomic: 9.7% decrease) (Figure 3a). Genetic correlations of SA within psychiatric diagnosis were similar to those of SA|MDD: of the 39 phenotypes with significant genetic correlation after Bonferroni correction, 21 phenotypes were in the smoking, education/cognition or socioeconomic risk categories (Figure 3b, Table S12).

**Figure 3:**
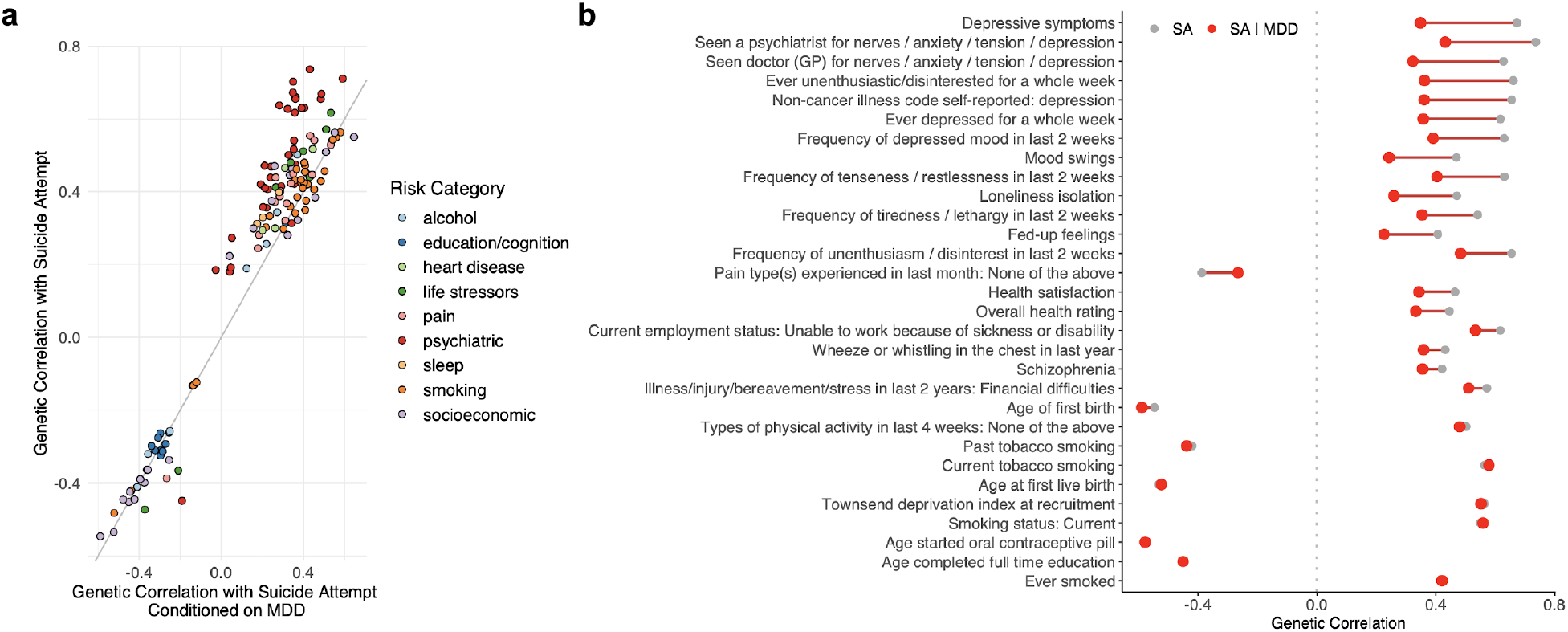
Conditioning suicide attempt on major depressive disorder reduces genetic correlation with psychiatric phenotypes but has limited effect on other traits. a) Comparison of significant genetic correlations with suicide attempt (SA) versus genetic correlations with SA conditioned on MDD (SA|MDD). Data include 133 significant genetic correlations after Bonferroni correction (P<0.05/768=6.51×10^−5^) annotated by risk category. b) Top 30 phenotypes with the most significant genetic correlations with SA before (in gray) and after conditioning on MDD (SA|MDD) (in red). Full genetic correlation results, including standard errors, are provided in Table S12.

## Discussion

We present a GWAS of suicide attempt in over 29,000 cases, identifying 2 genome-wide significant loci, including one locus more strongly associated with SA than with psychiatric disorders or other related traits. We demonstrate that a substantial proportion of the SNP-heritability of SA is independent of psychiatric diagnosis, by conditioning our GWAS results on the genetics of MDD and by examining the genetics of SA among individuals with a psychiatric diagnosis. Finally, we determine that the genetic liability to SA not mediated by psychiatric disorders is shared with the genetic architecture of traits related to smoking, socioeconomic traits, and poorer overall health.

The locus most strongly associated with SA was in an intergenic region on chromosome 7. The index SNP had a larger effect on SA than any common psychiatric disorder, remained genome-wide significant after conditioning on MDD and had a comparable effect size on SA within psychiatric diagnosis and self-harm ideation. Taken together, these results suggest that the genetic association with SA at this locus is not mediated through risk for psychiatric disorders. Current functional genomic data does not clearly link this variant to any gene, with the nearest gene being a long-non-coding RNA (*LINC01392*) located 149 kb away. The index SNP (rs62474683) is a methylation quantitative trait locus (mQTL), with the SA risk allele associated with decreased methylation of a nearby DNA methylation site (probe cg04544267) in blood^62^. However, this methylation site has not been linked to any gene transcript. Intriguingly, SA-risk alleles at this locus have previously been implicated at genome-wide significance in risk-taking behaviors^56^, smoking^57^, and insomnia^63^. While variants in the MHC also reached genome-wide significance for SA, this effect did not remain after conditioning the GWAS results on MDD. Indeed, variants in the MHC have previously been associated with risk for a range of psychiatric disorders including MDD^64^. This suggests that the association between the MHC and SA may be pleiotropic or potentially a byproduct of psychiatric diagnosis. Further investigation is needed to determine causality or direction for both of these loci.

Our GWAS results provide robust evidence of the 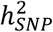 of SA, with an estimate of 6.8% on the liability scale (7.5% in the European-only subset). Importantly, conditioning on MDD resulted in a smaller but significant 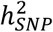 estimate (4.1%), which was on par with estimates from the GWAS of SA within psychiatric diagnosis (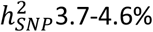 on the liability scale, using a prevalence of SA in psychiatric populations from 10-20%). These results corroborate previous reports^25,27^ of the independent genetic contribution to SA from genetic epidemiology studies and GWAS, and illustrate the importance of accounting for potential bias from the genetics of psychiatric disorders. Traditionally, GWAS of SA have sought to dissect this specific genetic component by conducting GWAS of SA within psychiatric diagnosis. More recently, a GWAS of SA in the iPSYCH Danish Registry took the approach of including a covariate for cases’ psychiatric diagnoses^27^. Here, we found complete genetic correlation between the GWAS of SA after conditioning on MDD and the GWAS of SA within psychiatric diagnosis (rg=1.13, SE=0.13), thus demonstrating that comparable results can be achieved via a statistical genetics approach. Since conditioning only requires GWAS summary statistics, this approach is readily applicable to different types of cohort and circumvents the need for samples with specific psychiatric diagnoses, detailed phenotypic information or individual-level genotype data available.

SA showed substantial positive genetic correlation with many psychiatric disorders, the highest being with MDD (rg=0.78, SE=0.03), consistent with previous reports^28,29,31^. Genetic overlap was also particularly strong with PTSD, ADHD, SCZ, and BIP (rg=0.44-0.74). After conditioning on MDD, there was a modest decrease in the genetic correlation of SA with most psychiatric disorders, but only significant decreases were observed with MDD, ASD, and self-harm ideation. Notably, after conditioning, SA was still strongly genetically correlated with MDD (rg=0.53, SE=0.06, P=8.85×10^−19^), representing pleiotropic effects between them. This genetic correlation would only be completely eliminated if all SNP effects on SA were mediated by MDD. Many studies have demonstrated extensive pleiotropy between psychiatric disorders^65,66^, and accordingly genetic overlap between SA and related disorders is anticipated. Our findings suggest that many pleiotropic genetic variants increase risk for SA directly, independent of their effects on psychiatric disorders. Examining the genetic liability to SA in a group of cases without psychiatric disorders would be a valuable future endeavor to corroborate these findings, however such individuals are a minority.

Genetic correlations were also examined between SA and 768 traits, with a focus on known risk factors and comorbidities. There was significant genetic correlation between SA and many other traits, including smoking, lower socioeconomic status, pain, lower educational attainment, reproductive traits, risk-taking behavior, sleep disturbances and poorer overall general health. While conditioning on MDD reduced the genetic correlations between SA and psychiatric disorders, in contrast, the genetic correlation of SA with most non-psychiatric traits remained unchanged. These results were largely corroborated using the GWAS of SA within psychiatric diagnosis, pointing to a consistent picture of shared genetic architecture between SA and these risk factors that is not a byproduct of psychiatric illness. There is substantial epidemiological literature on the relationship of risk factors including sleep disorders^12–15^, smoking^18–20^ and socioeconomic factors^67–69^ on SA, but less on the role of genetics. We have not assessed any causal role between the genetic risk of these traits and SA, but additional work on this topic will provide important insights and potentially highlight opportunities for risk stratification.

This first collaborative study by the International Suicide Genetics Consortium is almost 5-fold larger than any previous GWAS of SA, providing a substantial increase in statistical power. Furthermore, we have assessed the specificity of our findings to SA using two approaches. Nevertheless, several limitations must be acknowledged. Cases were defined across cohorts using a variety of diagnostic interviews, self-report, or hospital records, which may result in heterogeneity in the phenotype definition. Standard diagnostic criteria for SA are lacking and here sample sizes prohibited calculating genetic correlations across pairs of cohorts. Our GWAS included both cases of non-fatal SA and death by suicide which are imperfectly although highly genetically correlated (rg=0.77 between the University of Utah GWAS of suicide death and a meta-analysis of the remaining cohorts in our study). There is potential for misclassification of controls in the GWAS of SA within psychiatric diagnosis, as some patients may go on to make a suicide attempt later in life. We examined the genetic correlation between our GWAS of SA and psychiatric disorders, using publicly available GWAS summary statistics, however we note that the prevalence of SA amongst the cases in these GWAS are unknown. Finally, population, demographic and environmental factors are always present in genetic analyses and while our sample is large and diverse we did not have expansive data to stratify our analyses, to assess their possible contribution or confounding effects.

This work establishes the best-powered genetic analysis of SA to date. We identify SA risk loci and demonstrate a genetic component of SA that is not mediated through psychiatric disorders, but is shared with known risk factors. At present, PRS for SA do not have meaningful predictive utility and their premature use in either clinical or direct-to-consumer settings could be harmful. Dissecting the shared genetic architecture of SA, psychiatric disorders and other risk factors will be crucial to understanding the biological mechanisms of risk and assessing whether genetics can inform risk stratification or treatment.

## Data Availability

The policy of the International Suicide Genetics Consortium is to make genome-wide summary results public. Summary statistics will be made available online upon publication. This study included some publicly available datasets accessed through dbGaP - PGC bundle phs001254.v1.p1.

## Acknowledgements

We thank the participants who donated their time, life experiences and DNA to this research, and the clinical and scientific teams that worked with them. Statistical analyses were carried out on the NL Genetic Cluster Computer (http://www.geneticcluster.org) hosted by SURFsara and the Mount Sinai high performance computing cluster (http://hpc.mssm.edu), which is supported by the Office of Research Infrastructure of the National Institutes of Health under award numbers S10OD018522 and S10OD026880. This work was conducted in part using the resources of the Advanced Computing Center for Research and Education at Vanderbilt University, Nashville, TN. This work is supported by R01MH116269 (DMR) and R01MH121455 (DMR). Research reported in this publication was also supported by NIGMS of the National Institutes of Health under award number T32GM007347 (JK). The content is solely the responsibility of the authors and does not necessarily represent the official views of the National Institutes of Health.

